# Mental health outcomes and associations during the coronavirus disease 2019 pandemic: A cross-sectional survey of the US general population

**DOI:** 10.1101/2020.05.26.20114140

**Authors:** Bella Nichole Kantor, Jonathan Kantor

## Abstract

Pandemic coronavirus disease 2019 (COVID-19) may lead to significant mental health stresses, potentially with modifiable risk factors. To determine the presence of and magnitude of associations between baseline associations and anxiety and depression in the US general population, we performed an internet-based cross-sectional survey of an age-, sex-, and race-stratified representative sample from the US general population. Degrees of anxiety, depression, and loneliness were assessed using the 7-item Generalized Anxiety Disorder scale (GAD-7), the 9-item Patient Health Questionnaire (PHQ-9), and the 8-item UCLA Loneliness Scale, respectively. Unadjusted and multivariable logistic regression analyses were performed to determine associations with baseline demographic characteristics. A total of 1,005 finished surveys were returned of the 1,020 started, yielding a completion rate of 98.5% in the survey panel. The mean (SD) age of respondents was 45 (16), and 494 (48.8%) were male. Baseline demographic data were similar between those that were (n=663, 66.2%) and were not (n=339, 33.8%) under a shelter in place/ stay at home order, with the exception of sex and geographic location. Overall, 264 subjects (26.8%) met criteria for an anxiety disorder based on a GAD-7 cutoff of 10; a cutoff of 7 yielded 416 subjects (41.4%) meeting clinical criteria for anxiety. On multivariable analysis, male sex (OR 0.65, 95% CI [0.49, 0.87]) and living in a larger home (OR 0.46, 95% CI [0.24, 0.88]) were associated with a decreased odds of meeting anxiety criteria. Rural location (OR 1.39, 95% CI [1.03, 1.89]), loneliness (OR 4.92, 95% CI [3.18, 7.62]), and history of hospitalization (OR 2.04, 95% CI [1.38, 3.03]), were associated with increased odds of meeting anxiety criteria. 232 subjects (23.6%) met criteria for clinical depression. On multivariable analysis, male sex (OR 0.71, 95% CI [0.53, 0.95]), increased time outdoors (OR 0.51, 95% CI [0.29, 0.92]), and living in a larger home (OR 0.35, 95% CI [0.18, 0.69]), were associated with decreased odds of meeting depression criteria. Having lost a job (OR 1.64, 95% CI [1.05, 2.54]), loneliness (OR 10.42, 95% CI [6.26, 17.36]), and history of hospitalization (OR 2.42, 95% CI [1.62, 3.62]), were associated with an increased odds of meeting depression criteria. Income, media consumption, and religiosity were not associated with mental health outcomes. Anxiety and depression are common in the US general population in the context of the COVID-19 pandemic, and are associated with potentially modifiable factors.

---

The Coronavirus disease 2019 (COVID-19) pandemic has led to unprecedented levels of movement restriction, job losses, and economic uncertainty in the United States and around the world.^1^ Concerns regarding illness, death, and the death of loved ones may be compounded by financial uncertainty, as reports of mass unemployment with variable international governmental responses circulate.^2^

Mental health outcomes have been associated with pandemics in the past.^3-5^ While there has been a rapid response to the COVID-19 pandemic in terms of nonpharmaceutical interventions, vaccine development, and medical support, little comprehensive planning has been performed to predict and respond to the possible mental health crisis that could emerge from the pandemic, and the only data available on general public responses to the pandemic are in Chinese populations.^6,7^ These data are echoed by recent research that has suggested that healthcare workers have a significant burden of mental health challenges in the face of COVID-19.^8^ Moreover, pandemics and other natural disasters may disproportionately affect those with underlying mental illness.^9^

We therefore sought to investigate the prevalence of anxiety and depression in the general US population in the context of the early COVID-19 pandemic, and explore associations of these mental health outcomes with loneliness (of particular concern given enhanced social distancing and isolation), health status, socioeconomic status, residence size, time spent outdoors, and other baseline demographic characteristics. A better understanding of the prevalence of these mental health outcomes and their putative risk factors may help guide public policy in establishing improved guidelines for those required to stay at home.

## Methods

### Study Design

This study is a cross-sectional, internet-based survey performed via age, sex, and race stratification, conducted between March 29, 2020 and March 31, 2020. Responses to all survey questions were recorded (Supplemental file). This study was deemed exempt by the Ascension Health institutional review board.

We developed an online survey using the Qualtrics platform (Qualtrics Corp, Provo, Utah) after iterative online pilot testing. The survey was distributed to a representative sample of the US population using Prolific Academic (Oxford, United Kingdom), an established platform for academic survey research.^10^ Respondents were rewarded with a small payment (<US$1). Participants provided consent and were permitted to terminate the survey at any time. All surveys were anonymous and confidential, with linkages between data performed using a 24-character alphanumeric code. The investigators had no access to identifying information at any time.

### Participants

This internet-based survey was stratified by age, *sex*, and race to reflect the makeup of the general US population. Sample size calculations were conducted for the primary endpoint of detecting a 10% difference in the Generalized Anxiety Disorder-7 scale (GAD-7) between those that were and were not under a stay at home order at the time of survey completion. 682 subjects (341 per group) would be adequate to detect a 10% change in GAD-7 with 80% power and with an alpha of 0.05, assuming a baseline GAD-7 mean of 11.6 with a standard deviation of 5.4 and assuming equal group sizes.^11^ We inflated our sample size to 1,000 given that approximately 2/3 of the US was under stay at home orders at the time of survey initiation and given uncertainty regarding changes in those orders over the duration of the survey, as well as to permit subgroup analyses.

### Outcome Measures

Demographic information was self-reported by respondents. Responses to a battery of questions regarding attitudes to the COVID-19 pandemic, were collected using Likert scales.

For our main outcome measures, anxiety and depression, validated scales were used. Anxiety was assessed using the GAD-7, a validated self-report scale for anxiety, with scores ranging from 0 (no anxiety) to 21 (extreme anxiety). Prior psychometric research suggested cutoffs as 0-4 (no anxiety); 5-9 (mild anxiety); 10-14 (moderate anxiety); and 15-21 (severe anxiety).^8,11^

Depression was assessed with the Patient Health Questionnaire-9 (PHQ-9), a validated measure for clinical depression.^12^ Scores range from 0 (no depression) to 27 (severe depression). Prior psychometric research has suggested cutoffs as 0-4 (no depression); 5-9 (mild depression); 10-14 (moderate depression); and 15-27 (severe depression).^8,13^

Loneliness was quantified with the UCLA short-form loneliness scale (ULS-8), a validated measure of loneliness.^14^ Scores range from 8 (no loneliness) to 32 (extreme loneliness); no clinically meaningful cutoffs have been established psychometrically.

### Statistics

Normally distributed baseline demographic data are presented as mean values with 95% confidence intervals (CI). Outcomes that were not normally distributed are presented as medians with interquartile ranges (IQR). T-tests and chi-squared tests were used as appropriate for baseline continuous and categorical variables. Subgroup comparisons of non-normally distributed data were performed using the Kruskal Wallis test. Unadjusted and multivariable (adjusting for age and sex, which are not modifiable confounders) logistic regression odds ratios of association were assessed between the dependent variables of anxiety or depression, presented as dichotomous outcomes using the established cutoffs of 10 for both the GAD-7 and PHQ-9, and putative risk factors.

All statistical analyses were performed using Stata 13 for Mac (Stata Corporation, College Station, Texas).

## Results

### Baseline Characteristics

Of the 1,020 subjects who were recruited, 1,005 finished the survey, yielding a completion rate of 98.5%. The mean (SD) age of respondents was 45 (16), and 494 (48.8%) of the respondents were male; baseline respondent characteristics are outlined in Table 1. Baseline demographic data were similar between those that were (n=663, 66.2%) and were not (n=339, 33.8%) under a shelter in place or stay at home order, with the exception of sex and geographic location (urban versus rural status). The median (IQR) ULS-8 score for loneliness was 16 (8), similar to baseline estimates from previous studies.^14-16^

**Table 1.** Demographic and baseline characteristics of respondents, overall and by shelter in place status.

|  | No. (%) |  |  |
| --- | --- | --- | --- |
|  |  | *Under a shelter in place order* |  |
| Characteristic | Total | Yes | No |
| Overall | 1005 (100) | 681 (66.8) | 389 (33.2) |
| Sex* |  |  |  |
| Men | 494 (48.8) | 310 (46.1) | 184 (54.3) |
| Women | 518 (51.2) | 363 (53.9) | 155 (45.7) |
| Age, y |  |  |  |
| 18-30 | 250 (24.5) | 165 (24.2) | 85 (25.1) |
| 31-40 | 204 (20.0) | 139 (20.4) | 65 (19.2) |
| 41-50 | 146 (14.3) | 100 (14.7) | 46 (13.6) |
| 51-60 | 198 (19.4) | 130 (19.1) | 68 (20.1) |
| >60 | 222 (21.8) | 147 (21.6) | 75 (22.1) |
| Education level |  |  |  |
| < High school | 11 (1.1) | 8 (1.2) | 3 (0.9) |
| High school | 117 (11.7) | 67 (10.1) | 50 (14.8) |
| Some college | 228 (22.8) | 149 (22.4) | 79 (23.4) |
| Associates | 103 (10.3) | 66 (9.9) | 37 (11.0) |
| Bachelor's | 358 (35.7) | 246 (37.0) | 112 (33.2) |
| Graduate | 185 (18.5) | 129 (19.4) | 56 (16.6) |
| Employment status |  |  |  |
| Full time | 461 (45.2) | 303 (44.5) | 158 (46.6) |
| Part time | 170 (16.7) | 115 (16.9) | 55 (16.2) |
| Not employed | 389 (38.1) | 263 (38.6) | 127 (37.2) |
| Marital status |  |  |  |
| Married | 414 (40.6) | 273 (41.2) | 139 (41.0) |
| Unmarried | 606 (59.4) | 390 (58.8) | 200 (59.0) |
| Religious |  |  |  |
| Yes | 387 (37.9) | 252 (37.0) | 135 (39.8) |
| No | 543 (53.2) | 361 (53.0) | 182 (53.7) |
| Ambivalent | 90 (8.8) | 68 (10.0) | 22 (6.5) |
| Income |  |  |  |
| <\$10,000 | 167 (16.4) | 115 (16.9) | 52 (15.3) |
| \$10,000-\$30,000 | 234 (22.9) | 154 (22.6) | 80 (23.6) |
| \$30,001-\$50,000 | 220 (21.6) | 137 (20.1) | 83 (24.5) |
| \$50,001-\$80,000 | 201 (19.7) | 131 (19.2) | 70 (20.7) |
| \$80,001-\$100,000 | 63 (6.2) | 42 (6.2) | 21 (6.2) |
| \$100,001-\$150,000 | 91 (8.9) | 71 (10.4) | 20 (5.9) |
| >\$150,000 | 44 (4.3) | 31 (4.6) | 13 (3.8) |
| Location* |  |  |  |
| Urban | 743 (72.8) | 517 (75.9) | 226 (66.7) |
| Rural | 277 (27.2) | 164 (24.1) | 113 (33.3) |

All values are listed as number (%)
\* p<0.05.

### Anxiety

The median (IQR) GAD-7 score was 5 (9), and 513 subjects (52.1%) of subjects had at least mild anxiety. Overall, 264 subjects (26.8%) met criteria for an anxiety disorder based on a GAD-7 cutoff of 10 (Table 2). Adopting a more liberal GAD-7 cutoff of 7, as used in a recent study on healthcare worker anxiety in the COVID-19 context,^8^ would yield 416 subjects (41.4%) meeting clinical criteria for anxiety. Women (p=0.002) and those living in rural areas (p=0.041), reported more severe anxiety than men and those in urban areas, respectively.

**Table 2.** Anxiety and depression severity, by selected baseline characteristics. Abbreviations: GAD-7: Generalized Anxiety Disorders scale PHQ-9: Patient Health Questionnaire

|  | No. (%) |  |  |  |  |  |  |  |  |  |  |  |  |
| --- | --- | --- | --- | --- | --- | --- | --- | --- | --- | --- | --- | --- | --- |
|  |  | Sex |  |  | Shelter in place |  |  | Location |  |  | Married |  |  |
| Severity | Overall | Male | Female |  | Yes | No |  | Urban | Rural |  | Yes | No |  |
| **GAD-7** |  |  |  |  |  |  |  |  |  |  |  |  |  |
| None | 472<br>(47.9) | 260<br>(53.9) | 212<br>(42.2) | P=0.002 | 299 (45.9) | 172 (52.1) | P=0.314 | 337 (47.5) | 134 (49.5) | P=0.041 | 195 (48.2) | 277 (47.8) | P=0.894 |
| Mild | 249<br>(25.3) | 111<br>(23.0) | 138<br>(27.4) |  | 172 (26.4) | 76 (23.0) |  | 194 (27.4) | 53 (19.6) |  | 106 (27.2) | 143 (24.7) |  |
| Moderate | 132<br>(13.4) | 60<br>(12.5) | 72<br>(14.3) |  | 89 (13.7) | 42 (12.7) |  | 93 (13.1) | 39 (14.4) |  | 51 (12.6) | 81 (14.0) |  |
| Severe | 132<br>(13.4) | 51<br>(10.6) | 81<br>(16.1) |  | 92 (14.1) | 40 (12.1) |  | 85 (12.0) | 45 (16.6) |  | 53 (13.1) | 79 (13.6) |  |
| **PHQ-9** |  |  |  |  |  |  |  |  |  |  |  |  |  |
| None | 518<br>(52.7) | 278<br>(58.3) | 240<br>(47.4) | P=0.008 | 340 (52.2) | 176 (53.3) | P=0.457 | 373 (52.8) | 143 (52.6) | P=0.714 | 239 (59.9) | 279 (47.8) | P<0.0001 |
| Mild | 233<br>(23.7) | 102<br>(21.4) | 131<br>(25.9) |  | 149 (22.9) | 84 (25.5) |  | 165 (23.4) | 68 (25.0) |  | 92 (23.10) | 141 (24.1) |  |
| Moderate | 116<br>(11.8) | 48<br>(10.1) | 68<br>(13.4) |  | 84 (12.9) | 32 (9.7) |  | 114 (11.7) | 27 (9.9) |  | 34 (8.5) | 82 (14.0) |  |
| Severe | 116<br>(11.8) | 49<br>(10.3) | 67<br>(13.2) |  | 78 (12.0) | 38 (11.5) |  | 115 (11.8) | 34 (12.5) |  | 34 (8.5) | 82 (14.0) |  |

Unadjusted logistic regression analysis demonstrated that men were less likely to meet criteria for anxiety (OR 0.67, 95% CI [0.51, 0.89]) while those who lost their job (OR 1.61, 95% CI [1.45, 2.45]), had been hospitalized within the past 2 years (OR 1.86, 95% CI [1.27, 2.73]), or were in the most lonely quartile (OR 5.39, 95% CI [3.53, 8.24]), were more likely to meet criteria for anxiety. On multivariable analysis controlling for age and sex as confounders, male sex (OR 0.65, 95% CI [0.49, 0.87]), and living in a larger home (OR 0.46, 95% CI [0.24, 0.88]) were associated with a decreased odds of meeting anxiety criteria. Rural location (OR 1.39, 95% CI [1.03, 1.89]), loneliness (OR 4.92, 95% CI [3.18, 7.62]), and history of hospitalization within the past 2 years (OR 2.04, 95% CI [1.38, 3.03]), were independent risk factors for meeting anxiety criteria (Table 3
).

**Table 3.** Risk factors for anxiety and depression in unadjusted and multivariable analyses.

|  | Unadjusted OR (95% CI) | P value | Adjusted<sup>†</sup> OR (95% CI) | P value |
| --- | --- | --- | --- | --- |
| **GAD-7 (Anxiety)\*** |  |  |  |  |
| Sex |  |  |  |  |
| Male | 0.67 (0.51, 0.89) | 0.005 | 0.65 (0.49, 0.87) | 0.003 |
| Female | 1 [Reference] |  | 1 [Reference] |  |
| Location |  |  |  |  |
| Rural | 1.32 (0.98, 1.79) | 0.066 | 1.39 (1.03, 1.89) | 0.034 |
| Urban | 1 [Reference] |  | 1 [Reference] |  |
| Hospitalized in past 2 years |  |  |  |  |
| Yes | 1.86 (1.27, 2.73) | 0.001 | 2.04 (1.38, 3.03) | <0.0001 |
| No | 1 [Reference] |  | 1 [Reference] |  |
| Lost job due to COVID-19 |  |  |  |  |
| Yes | 1.61 (1.05, 2.45) | 0.028 | 1.53 (0.99, 2.35) | 0.055 |
| No | 1 [Reference] |  | 1 [Reference] |  |
| Loneliness (ULS-8)<sup>‡</sup> |  |  |  |  |
| Most lonely quartile | 5.39 (3.53, 8.24) | <0.0001 | 4.92 (3.18, 7.62) | <0.0001 |
| Least lonely quartile | 1 [Reference] |  | 1 [Reference] |  |
| Home size (square feet) |  |  |  |  |
| <500 | 1 [Reference] |  | 1 [Reference] |  |
| 500-750 | 0.59 (0.32, 1.12) | 0.107 | 0.60 (0.31, 1.14) | 0.120 |
| 750-1000 | 0.38 (0.20, 0.70) | 0.002 | 0.37 (0.20, 0.69) | 0.002 |
| 1000-1500 | 0.48 (0.27, 0.85) | 0.013 | 0.50 (0.28, 0.91) | 0.024 |
| 1500-2000 | 0.50 (0.27, 0.90) | 0.022 | 0.53 (0.29, 0.98) | 0.043 |
| >2000 | 0.39 (0.21, 0.74) | 0.004 | 0.46 (0.24, 0.88) | 0.019 |
| **PHQ-9 (Depression)\*** |  |  |  |  |
| Sex |  |  |  |  |
| Male | 0.73 (0.55, 0.98) | 0.034 | 0.71 (0.53, 0.95) | 0.023 |
| Female | 1 [Reference] |  | 1 [Reference] |  |
| Location |  |  |  |  |
| Rural | 0.90 (0.65, 1.24) | 0.515 | 0.94 (0.67, 1.30) | 0.700 |
| Urban | 1 [Reference] |  | 1 [Reference] |  |
| Hospitalized in past 2 years |  |  |  |  |
| Yes | 2.16 (1.47, 3.17) | <0.0001 | 2.42 (1.62, 3.62) | <0.0001 |
| No | 1 [Reference] |  | 1 [Reference] |  |
| Lost job due to COVID-19 |  |  |  |  |
| Yes | 1.74 (1.13, 2.67) | 0.011 | 1.64 (1.05, 2.54) | 0.029 |
| No | 1 [Reference] |  | 1 [Reference] |  |
| Loneliness (ULS-8)<sup>‡</sup> |  |  |  |  |
| Most lonely quartile | 11.90 (7.21, 19.65) | <0.0001 | 10.42 (6.26, 17.36) | <0.0001 |
| Least lonely quartile | 1 [Reference] |  | 1 [Reference] |  |
| Home size (square feet) |  |  |  |  |
| <500 | 1 [Reference] |  | 1 [Reference] |  |
| 500-750 | 0.50 (0.26, 0.94) | 0.033 | 0.50 (0.26, 0.96) | 0.037 |
| 750-10000 | 0.41 (0.22, 0.76) | 0.004 | 0.40 (0.22, 0.76) | 0.005 |
| 1000-1500 | 0.42 (0.23, 0.76) | 0.004 | 0.45 (0.25, 0.83) | 0.010 |
| 1500-2000 | 0.35 (0.19, 0.65) | 0.001 | 0.38 (0.20, 0.72) | 0.003 |
| >2000 | 0.29 (0.15, 0.56) | <0.0001 | 0.35 (0.18, 0.69) | 0.002 |
| Time spent outdoors in past 3 days (hours) |  |  |  |  |
| 0 | 1 [Reference] |  | 1 [Reference] |  |
| <1 | 0.77 (0.47, 1.27) | 0.302 | 0.80 (0.48, 1.34) | 0.398 |
| 1-3 | 0.55 (0.34, 0.88) | 0.014 | 0.58 (0.35, 0.95) | 0.031 |
| 3-5 | 0.45 (0.24, 0.82) | 0.010 | 0.53 (0.28, 0.98) | 0.044 |
| >5 | 0.45 (0.25, 0.79) | 0.006 | 0.51 (0.29, 0.92) | 0.025 |

\* Score cutoff of 10 used for classification.
† Adjusted for age and sex.
‡ Assessed using the UCLA Loneliness Scale
Abbreviations:
GAD-7: Generalized Anxiety Disorders scale
PHQ-9: Patient Health Questionnaire

### Depression

The median (IQR) PHQ-9 score was 4 (8), and 465 (47.3%) of subjects reported at least mild depression by screening (Table 2). A total of 232 subjects (23.6%) met criteria for clinical depression. Women (p=0.008) and unmarried subjects (p<0.0001) reported more severe depression than men and those who are married, respectively.

Unadjusted logistic regression analysis demonstrated that men were less likely to meet criteria for depression (OR 0.73, 95% CI [0.55, 0.98]), while those who lost their job (OR 1.74, 95% CI [1.13, 2.67]), had been hospitalized within the past 2 years (OR 2.16, 95% CI [1.47, 3.17]), or were in the most lonely quartile (OR 11.90, 95% CI [7.21, 19.65]), were more likely to meet criteria for depression. On multivariable analysis controlling for age and sex as confounders, male sex (OR 0.71, 95% CI [0.53, 0.95]), increased time outdoors (OR 0.51, 95% CI [0.29, 0.92]), and living in a larger home (OR 0.35, 95% CI [0.18, 0.69]) were associated with a decreased odds of meeting depression criteria. Having lost a job (OR 1.64, 95% CI [1.05, 2.54]), loneliness (OR 10.42, 95% CI [6.26, 17.36]), and history of hospitalization within the past 2 years (OR 2.42, 95% CI [1.62, 3.62]), were associated with meeting depression criteria (Table 3).

## Discussion

In this first study of general US population mental health during the COVID-19 pandemic, we found high baseline levels of both anxiety and depression, independent of living under a shelter in place or stay at home order. More than half (52.1%) of respondents had at least mild anxiety, and 47.3% of subjects had at least mild depressive symptoms. Adopting the cutoff of 7 on the GAD-7 score for anxiety, as used in a recent study on COVID-19, would yield 416 subjects (41.4%) meeting clinical criteria for anxiety. This high burden of mental health concerns in the general population in the pandemic context suggests the need for further study and consideration for intervention.

Living in a larger home was associated with a reduced risk of both anxiety and depression; this effect was seen despite the lack of any association between anxiety or depression and household income and persisted when including income and number of household members into a multivariable model. Similarly, we found that increased time spent outdoors correlated with a reduction in depression (but not anxiety) risk, and those that spent more than an hour a day outdoors had approximately half the risk of depression as those that spent no time outdoors. This association of depression with time outdoors echoes prior research on access to green space access and its impact on mental health.^17^ Our finding that both larger living space and increased time spent outdoors correlate with a reduction in mental health burden may have actionable implications for public health initiatives and decisions regarding access to outdoor recreation areas during stay at home or shelter in place orders.

History of hospitalization, a rough measure of overall health status, was associated with an increased risk of both anxiety and depression. This effect persisted even when controlling for age and history of anxiety and depression, respectively, suggesting that those with a poorer health status may be at increased risk of adverse mental health outcomes in the context of the COVID-19 pandemic.

Media consumption, measured by the number of hours spent watching or reading about the pandemic, was not associated with the presence of anxiety or depression. Similarly, we did not detect significant associations between likelihood of meeting criteria for anxiety or depression and household income or religiosity on adjusted multivariable analyses.

Notably, we found that less than half of respondents had *no* anxiety; that is, more than half of subjects reported a level of anxiety that would at least be classified as mild. Conversely, 13.4% of subjects demonstrated severe anxiety, a higher proportion than has been reported even in healthcare workers responding to pandemic COVID-19.^8^

Loneliness is an established risk factor for both anxiety and depression,^15,18^ and we found an approximately 5-to 10-fold increase in odds of anxiety and depression, respectively, with being in the highest loneliness quartile. As with those living in smaller homes with minimal access to the outdoors, loneliness can be seen as an independent risk factor for anxiety or depression in the context of the COVID-19 pandemic.

### Limitations

This study has several limitations. First, as with any survey-based research, its generalizability may be limited. We used Prolific Academic for survey distribution in order to maximize our generalizability to the general US population by using an age-, sex-, and race-stratified survey panel design. As with any survey data, however, the sample willing to participate may not fully reflect the population of interest. Second, our study took place during the early phase of the COVID-19 pandemic, when shelter in place and stay at home orders were only just beginning. If anything, however, this underestimates the prevalence of anxiety and depression as these outcomes would only be expected to increase as restrictions persist, and highlights that even the anticipation of such restrictions may present a stressor. Third, as with any survey study, response bias and social desirability bias may play a role, though the anonymous survey design may help mitigate these concerns. Fourth, while our study relied on validated scales wherever possible, some survey questions were the product of pilot testing alone, and therefore their methodology—while consistent with the survey development literature—has not been fully vetted. Finally, and importantly, this cross-sectional study that lacks a comparator group cannot establish causation; therefore, we do not know whether the associations we describe are truly clinical risk factors.

## Data Availability

Data are available from the corresponding author.

## Conclusions

In this first study of mental health outcomes in the US population during the COVID-19 pandemic, we found high rates of depression and anxiety, with the most profound mental health effects in women, those with a history of hospitalization over the past two years, those who were most lonely, and those living in smaller homes and (for depression) those spending the least time outdoors.

